# Directionality of neural activity in and out of the seizure onset zone in focal epilepsy

**DOI:** 10.1101/2024.08.10.24311802

**Authors:** Hamid Karimi-Rouzbahani, Aileen McGonigal

**Author notes:** Correspondence to: Hamid Karimi-Rouzbahani.

## Abstract

Epilepsy affects over 50 million people worldwide, with approximately 30% experiencing drug-resistant forms that may require surgical intervention. Accurate localisation of the epileptogenic zone (EZ) is crucial for effective treatment, but how best to use intracranial EEG data to delineate the EZ remains unclear. Previous studies have used the directionality of neural activities across the brain to investigate seizure dynamics and localise the EZ. However, the different connectivity measures used across studies have often provided inconsistent insights about the direction and the localisation power of signal flow as a biomarker for EZ localisation. In a data-driven approach, this study employs a large set of 13 distinct directed connectivity measures to evaluate neural activity flow in and out the seizure onset zone (SOZ) during interictal and ictal periods. These measures test the hypotheses of “sink SOZ” (SOZ dominantly receiving neural activities during interictal periods) and “source SOZ” (SOZ dominantly transmitting activities during ictal periods). While the results were different across connectivity measures, several measures consistently supported higher connectivity directed towards the SOZ in interictal periods and higher connectivity directed away during ictal period. Comparing six distinct metrics of node behaviour in the network, we found that SOZ separates itself from the rest of the network allowing for the metric of “*eccentricity”* to localise the SOZ more accurately than any other metrics including “*in strength”* and “*out strength”*. This introduced a novel biomarker for localising the SOZ, leveraging the discriminative power of directed connectivity measures in an explainable machine learning pipeline. By using a comprehensive, objective and data-driven approach, this study addresses previously unresolved questions on the direction of neural activities in seizure organisation, and sheds light on dynamics of interictal and ictal activity in focal epilepsy.

## Introduction

More than 50 million people worldwide have epilepsy (Kwan & Brodie, 2000), and in about 30% of them, anti-seizure medications cannot effectively control the disorder (Chen et al., 2018). In cases of focal epilepsy, where seizures originate from a specific part of one hemisphere, those with drug-resistant forms may undergo presurgical evaluations to identify seizure-generating areas. This often involves intracranial electroencephalography (EEG) to delineate the epileptogenic zone (EZ), considered to be the sites primarily responsible for generating seizures (Lagarde & Bartolomei, 2024). If the clinical risk-benefit analysis is favourable, the EZ can be surgically removed or disconnected through resection or laser ablation. Despite advancements in multimodal approaches like magnetic resonance imaging (MRI), electroencephalography (EEG) and positron emission tomography (PET) scans, and extensive clinical expertise, accurate localisation of the EZ remains challenging and can hinder achieving seizure freedom (Vakharia et al., 2018).

Quantification methods have shown significant potential in localising the EZ by analysing intracranial EEG signals (Bartolomei et al., 2017; Bernabei et al., 2023; Gentiletti et al., 2022; Grinenko et al., 2018; Karimi-Rouzbahani & McGonigal, 2024). These methods typically focus on either the interictal or ictal time windows. In the ictal window, the most common epileptiform activities include low voltage fast activity (LVFA), baseline slow wave shifts, rhythmic spikes/spike-waves, and preictal low frequency spiking which are more dominant in seizure onset zone (SOZ^1^), where seizures are thought to originate from (Bernabei et al., 2023). These features have been successfully extracted from signals and used for EZ localization in previous studies (Di Giacomo et al., 2024; Grinenko et al., 2018). In the interictal window, traditional epileptiform characteristics which are quantified include interictal spikes/discharges and high-frequency oscillations (HFOs), with ongoing debate about which is more effective and ultimately possible increased predictive power by measuring their co-occurrence (Roehri et al., 2018). While clinical observations and animal models suggest spatial overlap between interictal and ictal neural activities to a variable degree (Avoli et al., 2006), the temporal balance between interictal and ictal states may depend on the directionality of activities measured using functional connectivity properties (Gunnarsdottir et al., 2022; Lagarde, Roehri, Lambert, Trebuchon, et al., 2018). Beyond the abovementioned epileptogenic patterns of ictal (e.g., LVFA, baseline shifts) and interictal activity (e.g., spikes and HFOs) various other more complex and often nonlinear features have also been successful in localising the SOZ in both windows (Andrzejak et al., 2012; Mooij et al., 2020; Sato et al., 2019). In a recent work, we evaluated the performance of a large array of 34 distinct signal features in localising the EZ in both interictal and ictal windows. We showed that signal power and network-based connectivity features were among the most localising features and were among the most generalisable features across patients (Karimi-Rouzbahani & McGonigal, 2024).

While many traditional methods for EZ localization focused on univariate or single-channel signal activity, there has been a shift towards multivariate, multi-channel or network-based localisation (Gallagher et al., 2023; Gunnarsdottir et al., 2022; Johnson et al., 2023; Karimi-Rouzbahani et al., 2024; Lagarde, Roehri, Lambert, Trebuchon, et al., 2018; A. Li et al., 2018, 2021). This approach aligns with the understanding of epilepsy as a network disorder (Kramer & Cash, 2012; Spencer, 2002) and has demonstrated better localisation performance compared to univariate methods in several studies (Balatskaya et al., 2020a; Bernabei et al., 2022; Kini et al., 2019). These have established the connectivity measures as valuable biomarkers for EZ localisation. In the interictal period, the consensus is that connectivity is higher within the EZ than within the non-involved zones (NIZ) and that EZ is relatively disconnected from the NIZ (Johnson et al., 2023; Lagarde, Roehri, Lambert, Trébuchon, et al., 2018). In the ictal period, areas within the SOZ increase their internal connectivity and become less connected to non-SOZ areas upon seizure onset (Liu et al., 2021; Runfola et al., 2023; Schindler et al., 2007; Warren et al., 2010). While these studies showed consensus on increased connectivity within the EZ/SOZ and decreased connectivity between EZ/SOZ and non-involved areas, they generally focused on non-directed connectivity methods. Non-directed methods quantify the level of connectivity or interaction between areas but remain silent about the direction of activity flow. Specifically, they quantify the interaction without providing information about whether an area dominantly sends or receives neural activities from other connected areas.

Recent studies have focused on directed connectivity measures for the localisation of the EZ (Jiang et al., 2022; Z. Li et al., 2023; Nahvi et al., 2023). Directed connectivity methods provide insights into the direction of neural activities relative to each individual brain area and have shown better localising performance than non-directed connectivity measures (Narasimhan et al., 2020). The knowledge about the directionality of neural activity flows within and across epileptogenic networks can provide valuable insights such as where seizures are generated and how they are propagated.

Specifically, one hypothesis in epilepsy, which has attracted increasing attention, is the “interictal suppression hypothesis”, which posits that the EZ is inhibited by other brain areas in the interictal period (i.e. that this is why the brain does not continuously seize in subjects with epilepsy), and that seizures occur when this inhibition mechanism fails (Doss et al., 2024; Gunnarsdottir et al., 2022; Jiang et al., 2022; Johnson et al., 2023; Narasimhan et al., 2020; Paulo et al., 2022; Vlachos et al., 2017). However, findings are non-unanimous with some studies showing that interictal neural activities are dominantly towards the EZ (Gunnarsdottir et al., 2022; Jiang et al., 2022; Johnson et al., 2023; Narasimhan et al., 2020; Paulo et al., 2022; Vlachos et al., 2017) and others showing outward from the SOZ (Amini et al., 2011; Bettus et al., 2011; Lagarde, Roehri, Lambert, Trebuchon, et al., 2018; Wilke et al., 2009). In the ictal period also, the intuition is that SOZ not only initiates the seizures but also transmits activity to other brain areas. However, the application of directed connectivity measures has shown discrepant results with some studies showing that ictal activity propagates from the SOZ to other areas (Balatskaya et al., 2020b; Courtens et al., 2016; Jung et al., 2011; Yang et al., 2018) supporting a change of role from “sink” to “source” of activity (Gunnarsdottir et al., 2022; Jiang et al., 2022). Nonetheless, other studies have shown the opposite direction of neural activities dominantly *towards* the SOZ in the ictal period (An et al., 2020; Janca et al., 2021; Mao et al., 2016; Nahvi et al., 2023). While the dominant outflow from the SOZ can be explained by a significant increase in power in the SOZ being propagated to other areas (Liou et al., 2020), it has been postulated that the latter can possibly be justified by potential efforts of the non-SOZ areas to inhibit and stop the seizure (surrounding inhibition) (Schevon et al., 2012).

One main reason for the discrepancy between studies evaluating the direction of activity flow (connectivity) could be the different directed connectivity methods used in different studies (Lagarde et al., 2022; Lagarde & Bartolomei, 2024). As each of directed connectivity methods is mathematically distinct, they rely on distinct aspects of signals and their potential relationship (i.e., connectivity). For example, while directed transfer function (DTF) measures the influence of one signal on another in the frequency domain using power analysis and Fourier transform, directed coherence (DCOH) method uses a spectral transfer matrix and normalises the inflow from one signal to another by their noise covariance (Baccalá & Sameshima, 2001). Considering such significant mathematical difference in the directed connectivity methods, this could be suspected to potentially contribute different levels and sometimes opposite directions of activity flow across studies (Plomp et al., 2014). Therefore, the development of more objective and data-driven approaches are required to determine the direction of activity towards and away from the SOZ (Doss et al., 2024; Lagarde & Bartolomei, 2024).

This study aims to establish the direction of neural activity in and out of the SOZ, in an unbiased, data-driven, and objective fashion. To that end, we used a large set of 13 mathematically distinct methods for quantifying directed connectivity used in the literature in the interictal and ictal periods. We then used network-analysis metrics (also known as connectomes (Doss et al., 2024)) including *in strength* and *out strength* to determine the dominant direction of broadband activity flow in and out of the SOZ. The aim is to see if the activities generally and dominantly flow towards or away from the SOZ and test the sink/source hypotheses in epilepsy with minimal effect of subjective method selection. Moreover, the knowledge about the directionality of neural activities in the interictal and ictal periods, if meaningful, can inform the development of automated EZ localisation methods.

Specifically, if the SOZ were consistently at the receiving end of activity in the interictal period, this could be a valuable localising piece of information for localisation algorithms. Therefore, to evaluate the localisation power of the directed connectivity methods in intracranial recording, we combined all directed connectivity methods to localise the SOZ in both interictal and ictal periods.

## Methods

### Dataset

This study uses a well-structured open-access intracranial dataset which brings together data from multiple centres (Bernabei et al., 2022; Kini et al., 2019). The dataset includes 57 patients who had been implanted with either subdural grid/strip (termed “electrocorticography” (ECoG)) or SEEG as their presurgical workup, and subsequently treated with surgical resection or laser ablation. Two patients’ data were excluded from our analyses as one had no interictal and the other no ictal recordings. Among the 55 patients analysed, 27 patients’ magnetic resonance imaging findings were lesional (28 non-lesional) and 35 patients were implanted with SEEG (20 ECoG (long-term subdural grid/strip recordings)). Thirty-four patients had Engel I, 6 Engel II, 11 Engel III and 2 had Engel IV outcomes. Resections/ablations targeted frontal (FRT) areas in 10 patients, temporal (TPR) in 24, mesiotemporal (MTL) in 15, insular in 2, frontoparietal in 1, parietal in 1 and mesiofrontal areas in 2 patients (see patient demographics in Supplementary Table 1). Clinically determined seizure onset channels were provided. Each patient had 2 interictal recordings and between 1 to 5 (*mean =* 3.7) ictal recordings/seizures (110 interictal and 204 ictal recordings over all patients). The interictal data was selected from awake brain activities determined both by the selection of day-time recordings (8 am – 8 pm) and the use of a custom non-REM sleep detector (explained in detail in (Bernabei et al., 2022)). The interictal data were at least 2 hours before the beginning of a seizure and at least 2 hours after a subclinical seizure, 6 hours after a focal seizure and 12 hours after a generalised seizure, free of spikes if possible and not within the first 72 h of recording to minimize immediate implant and anaesthesia effects. Epileptogenic zones/resected areas ranged from frontal, frontoparietal, mesiofrontal, temporal, mesiotemporal, parietal and insular areas.

### Pre-processing

Bad channels, as marked in the dataset, were excluded from analyses. An average of 105.6 contacts (*std* = 38.04) per patient remained after bad channels were removed. There was an average of 114.2 (*std* = 41.2) and 88.8 (*std* = 25.3) channels recorded in patients implanted with SEEG and ECoG, respectively. Among these, an average of 12.87% (*std* = 11.1%) of channels were in the SOZ area in each patient. The sampling frequencies of the signals varied across patients and ranged from 256 to 1024 Hz. We adjusted the sampling rate to 256 Hz across patients for analyses. We applied a 60Hz notch filter to the data to remove line noise. To reduce the computational load, we only kept a maximum of 30 contacts per patient for analysis - in a random sampling procedure, we kept all the channels within the SOZ, and the other channels (remaining of 30) were randomly selected from non-SOZ contacts which were inside the grey matter and at least 10 mm away from other contacts.

### Calculation of directed connectivity measures

We selected a 2-minute window of signal from each interictal recording (4 minutes per patient) and a patient-specific length of signal from each ictal recording (from seizure onset to the termination of seizure). Within those windows, we selected three 2-second epochs of data for analysis. The three epochs were chosen to capture early, mid and late dynamics of the signals within each recording.

Specifically, in the 2-minute interictal window, the early, mid and late epochs were separated by 59 seconds, and in the ictal period, the early and late epochs were separated by the length of the seizure while the mid epoch was in between the two windows. Our choice of 2-second epochs was to to select a mid-range epoch compared to previous studies which have used a wide range of epochs from 0.25s to 10 minutes to calculate connectivity in the interictal (Balatskaya et al., 2020b; Mooij et al., 2020; Sato et al., 2019) and from 20 to 60 seconds in the ictal (Li et al., 2018; Runfola et al., 2023) data.

We used the open-source python toolbox called PySpi (Cliff et al., 2023) which implements the largest set of directed and non-directed connectivity measures for time series (here intracranial EEG channels). We used the 13 available directed connectivity measures implemented in the toolbox for this work to follow an unbiased data-driven approach in analysis. The measures are categorised here into “information theory” (n=6), “frequency-domain” (n=6) and “time-domain” (n=1) methods. We briefly explain the characteristics of each connectivity measure below. For more information about each measure, the reader is advised to study the references cited for each measure.

#### Information theory measures

##### Additive Noise Model (ANM)

This measure assesses directed nonlinear dependence (or causality) of x → y under the assumption that the effect variable, y, is a function of a cause variable, x, along with an independent noise term (Hoyer et al., 2008). PySpi utilises the statistic from Causal Discovery Toolbox (CDT) as connectivity. This involves initially predicting y from x using a Gaussian process using a radial basis function kernel, followed by computing the normalized Hilbert-Schmidt Independence Criterion (HSIC) test statistic from the residuals. ANM is commonly used in causal inference studies, particularly when the underlying causal mechanisms are assumed to be deterministic and additive in nature.

##### Information-Geometric Causal Inference (IGCI)

This measure infers causal influence from x to y within deterministic systems featuring invertible functions (Janzing et al., 2012). In IGCI, causal inference is approached by examining the geometric structure of the joint probability distribution of variables. Specifically, IGCI focuses on estimating the causal influence of one variable (the cause) on another variable (the effect) by analysing the statistical dependencies between them. PySpi utilises CDT, where the difference in differential entropies is computed, with probability density estimated via nearest-neighbour estimators. One of the key features of IGCI is its ability to handle both linear and nonlinear causal relationships, making it applicable to a wide range of data types and systems. Additionally, IGCI can be used to infer causal relationships in scenarios where traditional statistical methods may not be suitable, such as when dealing with high-dimensional or noisy data.

##### Conditional Distribution Similarity Fit (CDS)

This measure provides a quantitative measure of the conditional relationship between variables and can help identify patterns of dependency or causality in empirical data (Cliff et al., 2023). It represents the standard deviation of the conditional probability distribution of y given x. This involves estimating the conditional probability distributions by discretising the values of the x and y and then computing the standard deviation of these conditional distributions. CDSF does not rely on specific parametric models to describe the relationship between variables, which makes it ideal for objective analyses.

##### Regression Error-Based Causal Inference (RECI)

This provides an assessment of the causal impact of x → y by measuring the error in a regression of y on x using a monomial (power product) model (Blöbaum et al., 2018). The rationale behind this method is that if there is a causal relationship from x to y, the regression model should capture most of the variation in y. This statistic corresponds to the Mean Squared Error (MSE) resulting from the linear regression of the cubic (with a constant term) of x with y. While linear regression models are commonly used, RECI can also be extended to handle nonlinear relationships between variables.

##### Causally Conditioned Entropy (CCE)

This measure quantifies the remaining uncertainty in time series y given the entire causal past of both time series x and y (Cliff et al., 2023). It is computed as a sum of conditional entropies of y given the past of both x and y with increasing history lengths. CCE is a sophisticated measure for assessing the causal influence of one time series on another by quantifying the remaining uncertainty in the target series after conditioning on the past values of both series. It is a versatile tool that can handle both linear and nonlinear dependencies. For computational efficiency, PySpi sets the history length of 10. This implies that the joint process is assumed to be, at most, a 10th-order Markov chain. We used a Gaussian kernel in this work.

##### Directed Information (DI)

It is a measure for assessing the information flow from a source time series x to a target time series y (Massey, 1990). It is calculated as the difference between the conditional entropy of y given its own past and the *CCE*. This measure provides an interpretable framework for understanding causal influence, as it directly quantifies the amount of information transferred from the source to the target time series. As in *CCE*, the computation of *directed information* is limited to a history length of 10. We used a Gaussian kernel in this work.

#### Frequency-domain measures

We also calculated several measures of directed connectivity in the frequency domain. For the frequency-domain measures, we used the full frequency range of 0 to 128Hz (the upper bound is limited by the Nyquist theorem to half the sampling rate) to obtain a broad-band index rather than focusing on a narrow frequency band. This provides general and objective results. The measures are calculated over 125 uniformly sampled bins across the 0 to 128Hz frequency range and averaged.

##### Group Delay (GD)

Group delay quantifies a directed, average time delay between two signals by assessing the slope of the phase differences as a function of frequency (derived through linear regression) (Hannan & Thomson, 1973). This slope is computed solely for coherence values that are statistically significant, and the time delay is acquired through a straightforward rescaling of the slope by 2π. The implementation provides the output in the form of the rescaled time delay statistic. GD’s ability to provide directional, frequency-dependent, and time-resolved measures of the interactions between signals is valuable. Its reliance on phase differences makes it particularly effective for studying the temporal dynamics of complex systems.

##### Phase Slope Index (PSI)

This measure serves as a directed metric for assessing information flow, computed using the complex-valued coherence (Nolte et al., 2008). Specifically, it evaluates the consistency of phase difference alterations across a predefined frequency range, with coherence acting as a weighting factor. The implementation computes the measure in the frequency domain. Its reliance on phase coherence makes it robust to noise and effective in identifying directed connectivity within specific frequency bands. However, *PSI* primarily assumes linear relationships in the phase domain. Nonlinear interactions may not be fully captured by this measure. Moreover, accurate phase difference estimation requires high-quality signals. Pre-processing steps such as filtering and artefact removal are crucial for reliable *PSI* computation.

##### Directed Transfer Function (DTF)

This measure uses cross-spectral density matrix which can be decomposed into a noise covariance matrix and a spectral transfer matrix (Eichler, 2006). The *directed transfer function* is derived from this decomposition to quantify the inflow from x to y. This inflow is normalized by the total inflow from all other signals into y, represented by the row-wise sum of the spectral transfer matrix. It can provide frequency-specific insights into connectivity, and its normalisation allows for direct comparison of connectivity between distinct pairs of signals. However, the computationally intensive nature of the method, its assumption linearity limits the application of DTF in detecting nonlinear relationships.

##### Directed Coherence (DCOH)

It is calculated from the inflow from x to y using the spectral transfer matrix (as described in DTF) and is then normalised by their noise covariance (Baccalá & Sameshima, 2001). It can provide frequency-specific insights into connectivity, and its normalisation allows for direct comparison of connectivity between distinct pairs of signals. However, the computationally intensive nature of the method, its assumption linearity limits the application of DCOH in detecting nonlinear relationships. Accurate estimation of *DCOH* requires high-quality signals. Noise and artefacts in the data can affect the reliability of the results, necessitating careful pre-processing steps.

##### Partial Directed Coherence (PDCOH)

The partial directed coherence from x to y is determined by the inflow (as described in DTF), normalized by the total outflow from all other signals into y (the column-wise sum of the spectral transfer matrix) (Baccalá & Sameshima, 2001). As an advantage to *DCOH*, by considering the influence of all other signals in the network (on the signals being evaluated), *PDCOH* provides a more accurate assessment of the true directional relationships between specific signal pairs.

##### Spectral Granger Causality (SGC)

This measure extends the concept of Granger Causality to the frequency domain, enabling the assessment of causal interactions between signals at specific frequencies (Friston et al., 2014). It is calculated using the spectral transfer matrix and noise covariance. These are estimated through either a parametric (VAR model) approach or a nonparametric (spectral factorization) approach. We used the *nonparametric* method to minimise subjective parameter settings. In the PySpi toolbox is implemented using the Spectral Connectivity Toolbox. *SGC’s* ability to provide frequency-specific and normalized measures of causality makes it particularly useful where understanding the dynamics of complex systems is important. However, its reliance on linearity and sensitivity to signal quality should be considered when interpreting the results.

#### Time-domain measure

##### Linear Model Fit (LMFIT)

Linear regression is a widely employed method for assessing independence via model fittings (Cliff et al., 2023). We employed ridge regression from the toolbox, which uses ℓ2-norm regularization and the mean squared error (MSE) resulting from a regression of y on x. This measure is a powerful and widely used statistical method for modelling the directed relationship between signals. Its simplicity, interpretability, and efficiency make it a valuable tool for relationship analysis, and trend analysis in various fields of study. However, it has several limiting assumptions including that observations are independent of each other. Violation of this assumption (e.g., autocorrelation in time series data) can lead to inaccurate estimates and predictions. Linear regression assumes a linear relationship between signals. If the true relationship is nonlinear, the model may provide biased or inaccurate results. Finally, overfitting can occur when the model is too complex relative to the amount of data available. This can result in poor generalization to new data and unreliable predictions.

### Node metrics

To characterise the role and behaviour of each node in the brain network, we used several network analysis metrics (i.e., connectomics). Specifically, in network analysis, each network is composed of **nodes** which are the electrode contacts here and **links** which are the (assumed) inter-node connections. Using the open-source brain connectivity toolbox (Rubinov & Sporns, 2010), we extracted six node metrics to evaluate the node behaviour in the network:

*In strength* is the sum of inward link weights (connectivity values). Nodes with higher *in strength* are influential receivers within the network, as they accumulate a significant amount of incoming influence, resources, or interactions from other nodes.

*Out strength* is the sum of outward link weights. Nodes with higher *out strength* are influential senders within the network, as they contribute a significant amount of outgoing influence, resources, or interactions to other nodes.

*First passage time* is the expected number of steps it takes a random walker to reach one node from another. Nodes with higher first passage times are often located on the periphery, farther away from central or densely connected regions.

*Clustering coefficient* is the fraction of triangles around a node and is equivalent to the fraction of node’s neighbours that are neighbours of each other. Nodes with higher clustering coefficients are typically located in densely connected neighbourhoods. These nodes have many connections to neighbouring nodes, forming cohesive groups or communities.

*Eccentricity* is the maximal shortest path length between a node and any other node. Nodes with higher eccentricity are typically located on the periphery of the network. They are farther away from the central core or densely connected regions.

*Betweenness centrality* is the fraction of all shortest paths in the network that contain a given node. Nodes with higher betweenness centrality often serve as bridges or connectors between different clusters, communities, or groups within the network. They lie on many of the shortest paths connecting nodes in different regions. Nodes with lower betweenness centrality are typically located on the periphery of clusters or communities within the network. They have fewer connections to other nodes and are less likely to lie on shortest paths between nodes.

### Multivariate pattern classification for SOZ localisation

We employed a standard multivariate pattern classification approach to localize the seizure onset zone (SOZ), distinguishing contacts within the SOZ from those outside it (non-SOZ) (Karimi- Rouzbahani & McGonigal, 2024). Initially, we computed inter-channel directed connectivity values. Subsequently, we calculated the six aforementioned node metrics for each contact based on the connectivity matrix, which has a size of N×N, where N represents the number of nodes or contacts. These metrics were then concatenated and utilised as features for the classifiers. The classification performance gauged the discriminability of SOZ and non-SOZ contacts using directed connectivity measures, assessed by the area-under-the-curve (AUC) metric for comprehensive, threshold-free classification performance. Consistent with recent localisation studies (Jiang et al., 2022; Karimi-Rouzbahani & McGonigal, 2024), we employed decision tree (DT) classifiers, treating each contact as an observation in classification. Our DT classifiers utilised a random forest algorithm with 50 bags of feature combinations, suitable for nonlinear feature classifications and offering insights into feature contributions. This approach clarifies the “contribution” of each feature by permuting the contact labels (i.e., SOZ vs. non-SOZ) in each feature separately and assessing its impact on performance, where contribution is inversely proportional to performance drop. For classification, we conducted separate analyses within interictal and ictal time windows for each patient, employing a 10-fold cross-validation procedure. This procedure was applied individually for each recording data from ictal and interictal packets and also their combinations. To address the imbalance in the number of SOZ to non-SOZ contacts and prevent bias toward one class in classification, we employed an up-sampling procedure to increase the number of observations for the class with fewer observations/contacts, repeating each classification of data 1000 times before averaging the results. Additionally, we generated chance-level performances by shuffling (SOZ/non-SOZ) contact labels 1000 times and recalculating the classification performance, resulting in 1000 chance-level classification outcomes against which we assessed the significance of our true classification performances.

### Statistical analysis

We employed Bayes Factor (BF) analysis for statistical inference. We compared the AUC levels against chance-level AUCs and assessed main effects on classifications. We interpreted levels of BF evidence strictly: BFs above 10 and below 1/10, were considered evidence for the alternative and null hypotheses, respectively. BFs falling between 1/10 and 10 were regarded as providing insufficient evidence either way, indicating that no conclusions could be drawn about the difference between a pair of variables.

To evaluate the evidence for the null and alternative hypotheses regarding at-chance and above-chance classification, respectively, we compared the classification rates in each analysis with those obtained from null distributions of the same analysis. For this purpose, we conducted an unpaired Bayes factor t-test for the alternative (i.e., difference from chance; H1) and the null (i.e., no difference from chance; H0) hypotheses.

To assess the evidence for the null and alternative hypotheses regarding the difference between contributions across measures and metrics, we compared the contributions obtained from each of these conditions using paired Bayes factor t-tests.

For evaluating the main effects of surgery outcome (Engel I/Engel II-IV), region of resection (FRT/TPRMTL), pathology (lesional/non-lesional), and recording modality (SEEG/ECoG), we employed a Bayes factor ANOVA. In this analysis, these four factors served as independent variables, with classification/generalization AUC as the dependent variable.

To ensure statistical power in ANOVA, we excluded patients with insular, frontoparietal, parietal, and mesiofrontal resections, where the sample size was less than 3. Priors for all Bayes factor analyses were determined based on Jeffrey-Zellner-Siow priors (Jeffreys, 1998; Zellner & Siow, 1980), which are derived from the Cauchy distribution based on the effect size initially calculated in the algorithm using t-tests (Rouder et al., 2012).

### Data and code availability

The dataset used in this study was from previous studies and is available at https://openneuro.org/datasets/ds004100/versions/1.1.3. The code developed for this project is available at https://github.com/HamidKarimi-Rouzbahani/Intracranial_epilepsy_connectivity.

## Results

We analysed directed connectivity measures of intracranial neural signals in epileptic patients towards two goals. First, we tested the sink-source hypotheses which suggest that SOZ areas dominantly receive neural activities during the interictal period (i.e., confirming “sink SOZ” hypothesis; Figure 1A). This direction was hypothesised to reverse in the ictal period (i.e., confirming “source SOZ” hypothesis; Figure 1A). Second, we tested to see if directed connectivity measures could localise the SOZ (Figure 1D). Importantly, to address these goals in a data-driven and objective manner, we used a large set of 13 distinct measures of directed connectivity to analyse the data which can clarify if the difference between connectivity methods used could explain the discrepancy in the literature.

**Figure 1.**
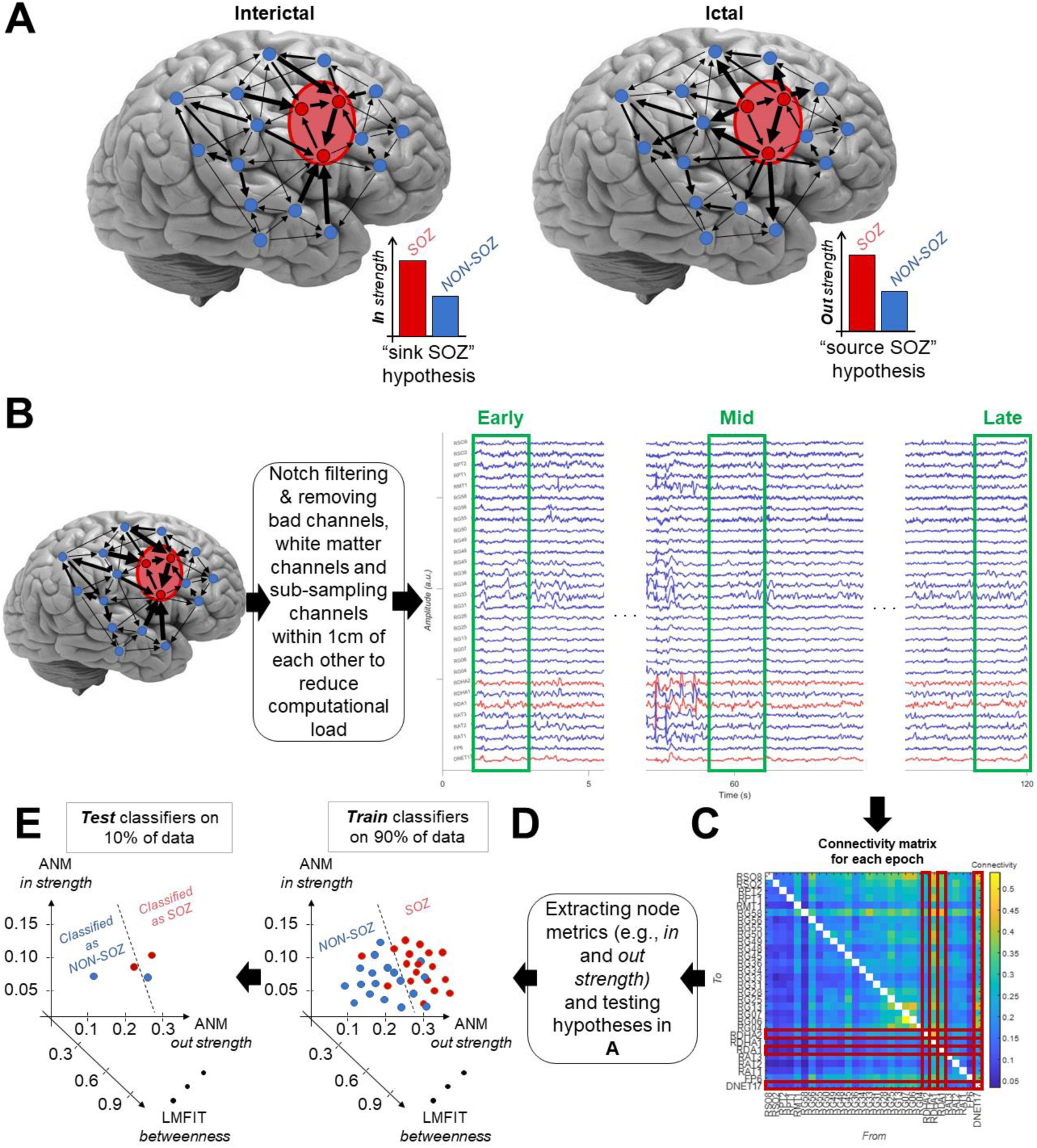
Hypotheses and the proposed localisation pipeline. **(A)** Sample implanted electrodes where seizure onset zone (SOZ, red nodes) shows dominantly outgoing activities compared to non-SOZ (blue nodes), with arrows indicating the strength of activity flows/connectivity (thicker arrows representing stronger connections) predominantly from non-SOZ to SOZ between seizures (interictal period). The “SOZ sink” hypothesis suggests a higher *in strength* for SOZ than non-SOZ contacts in the interictal period and the “SOZ source” hypothesis suggests a higher *out strength* in SOZ than non-SOZ contacts in the ictal period. **(B)** Recorded signals are pre-processed, and three 2-second epochs of data are used in analyses. **(C)** An inter-contact directed connectivity matrix reflecting connectivity strengths (colour-coded) and direction (columns and rows represent source and destination areas, respectively), with red squares indicating SOZ contacts. **(D)** Several node metrics are extracted from the connectivity matrices to assess each node’s behaviour in the network. **(E)** Machine learning classifiers are trained to distinguish contacts within and outside the SOZ in a 10-fold cross-validation process.

### Strength of neural activity flow in and out of the SOZ

We evaluated the strength of neural activity towards and away from every node (electrode contact) using *in* and *out strength* node metrics, respectively. Accordingly, nodes with higher *in strength* have stronger inward connectivity than nodes with lower *in strength*, and nodes with higher *out strength* have stronger outward connectivity than nodes with lower *out strength* (see *methods*).

In the interictal period, across the 13 directed connectivity measures tested, there was evidence (BF >10) for higher *in strength* in SOZ than non-SOZ areas for 5 connectivity measures (ANM, DI, DTF, DCOH and PDCOH) and there was evidence (BF > 10) for higher *in strength* in non-SOZ than SOZ areas only for the CDS connectivity measure. There was insufficient evidence (0.1 < BF < 10) either way for the rest of the connectivity measures (Figure 2A). In the ictal period, there was evidence (BF >10) for higher *out strength* in SOZ than non-SOZ areas for ANM and SGC connectivity measures, respectively. There was insufficient evidence (0.1 < BF < 10) either way for the rest of the connectivity measures (Figure 2B).

**Figure 2.**
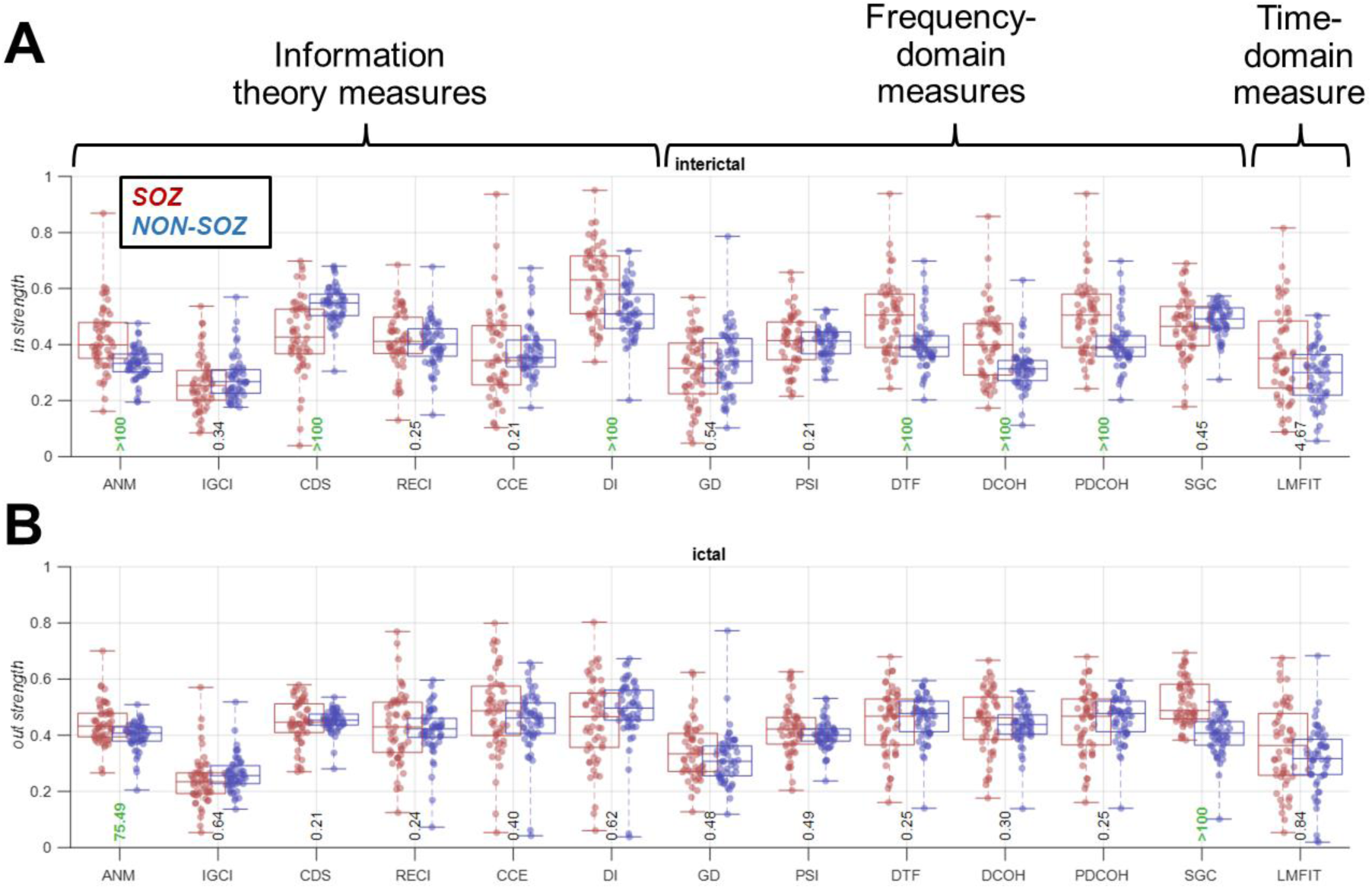
*In strength* in the interictal (A) and *out strength* in the ictal (B) period across connectivity measures. Connectivity measures are categorised into information theory, frequency-domain, and time-domain measures. Results are separated for the SOZ (red) and non-SOZ (blue) contacts with each dot showing data for one patient. Box plots show the distribution of data, its quartiles and median and whiskers indicate the maximum and minimum of the data across patients. Each dot indicates the data from one patient. Numbers below the bars indicate Bayesian evidence (BF > 10 indicated in green) for the difference between SOZ and non-SOZ data.

The results above were obtained by averaging the results from *early*, *mid* and *late* epochs (time windows) of the interictal and ictal periods (Figure 1B). To evaluate potential temporal variability in connectivity, we also evaluated *in strength* and *out strength* for each individual 2-second epoch (Supplementary Figure 1). The patterns of *in strength* were relatively similar across the three interictal windows (c.f., Figure 2A). The patterns of *out strength* were also similar across the three ictal windows and resembled the averaged results (c.f., Figure 2B).

While higher *in strength* in SOZ than non-SOZ areas (e.g., in the interictal period; c.f., Figure 2A) does not necessarily correspond to higher *out strength* in non-SOZ than SOZ areas, we tested this opposite non-hypothesised effect as well to ensure we are not overlooking a relevant effect (Supplementary Figure 2). In the interictal period, there was evidence (BF > 10) for higher *out strength* in non-SOZ than SOZ areas for the CDS connectivity measure only. However, there was evidence (BF > 10) respectively for higher *out strength* in SOZ than non-SOZ areas for ANM and SGC, respectively. In the ictal period, there was evidence (BF > 10) for higher *in strength* in non-SOZ than SOZ areas for the CDS only, but also evidence (BF > 10) for higher *in strength* in SOZ than non-SOZ areas. Therefore, the *out strength* during interictal period and the *in strength* during the ictal period provided inconsistent results across connectivity measures to support clear directions of activity flows.

Together, these results show that distinct measures of connectivity show variable results. Nonetheless, 5 out of 13 connectivity measures consistently supported the “sink SOZ” hypothesis in the interictal period. Similarly, but less strongly than in the interictal period, two connectivity measures supported the “source SOZ” hypothesis in the ictal period. These results were relatively stable within the interictal and ictal periods. Among the 13 connectivity measures evaluated, only the ANM measure supported both hypotheses.

### Switching of activity direction from the interictal to ictal period

The above results suggested that SOZ areas tend to be the receivers (i.e., sinks) in the interictal period and the transmitters (i.e., sources) of neural activity in the ictal period. However, it remains unclear if this tendency of switching roles between being a sink or source is consistent across patients (Doss et al., 2024). Specifically, the higher interictal *in strength* in SOZ compared to non-SOZ and the higher ictal *out strength* in non-SOZ compared to SOZ could have come from distinct subset of patients. To rule this out, we evaluated the correlation between effect sizes in the interictal and ictal periods: effects sizes were calculated as Δ = *in Strength_SOZ_* − *in Strength_non–SOZ_* during interictal and Δ = *out Strength_SOZ_* − *out Strength_non–SOZ_* during ictal period (Figure 3A). Majority (8 out of 13) of connectivity measures showed a positive correlation between the direction of effects across the interictal and ictal periods over patients with 5 reaching significance at p < 0.01 (*Pearson* correlation). The rest of the 5 connectivity measures showed non-significant negative correlations. Significant correlations suggest that patients in whom *in strength* was higher for SOZ compared to non-SOZ in the ictal period were the same patients in whom the *out strength* was also higher for non-SOZ compared to SOZ in the interictal period.

**Figure 3.**
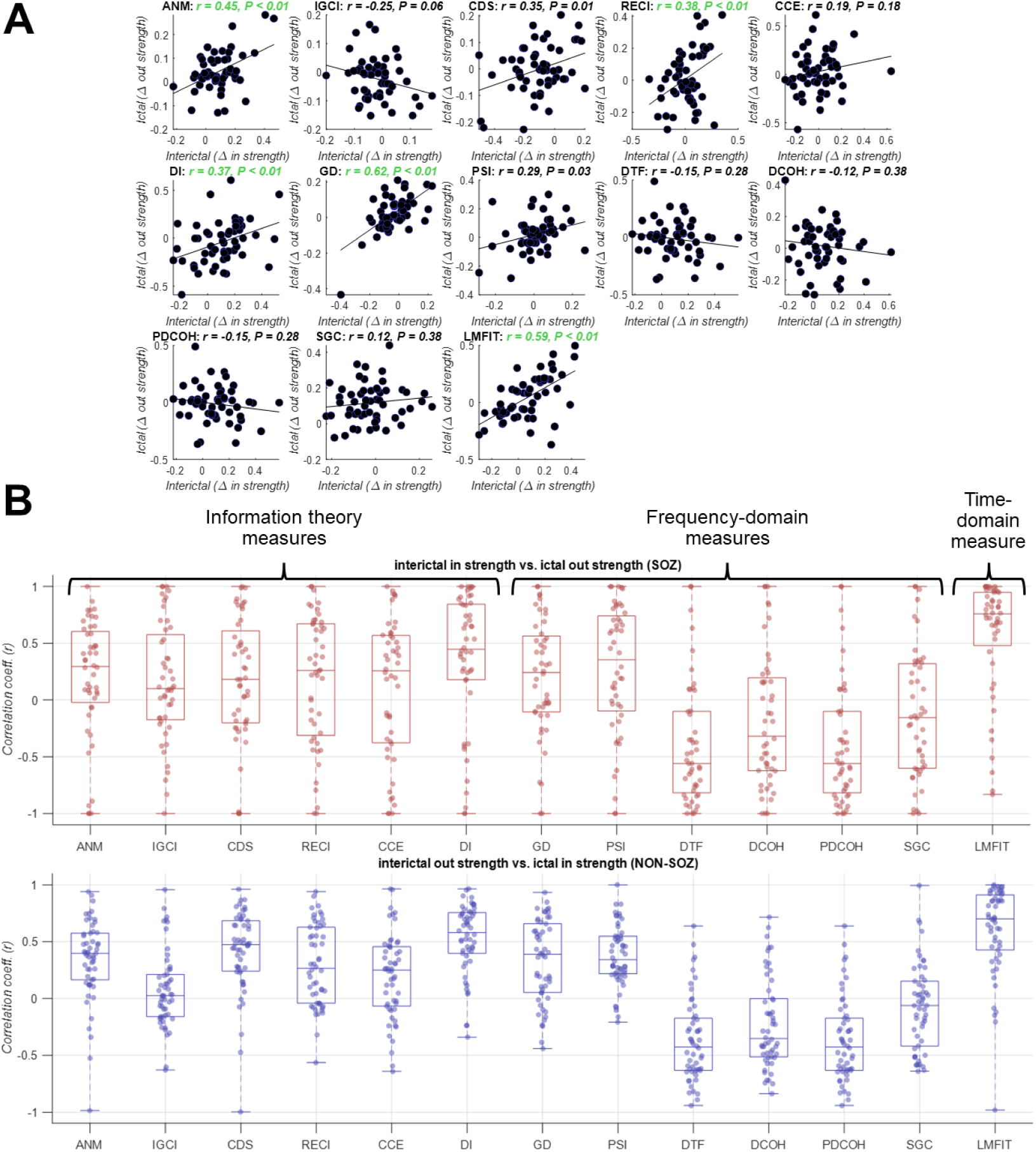
Correlation between interictal *in strength* and ictal *out strength*. **(A)** *Pearson* linear correlations between interictal and ictal effect sizes across patients with each dot showing data from one patient (i.e., effect size = difference between SOZ and non-SOZ contacts in terms of *in strength* in the interictal and *out strength* in the ictal period). Correlations and the corresponding p values are shown on top of each panel (significant results green: P < 0.01) with the slant line showing the best linear fit to the data. **(B)** *Pearson* linear correlation between interictal and ictal effect sizes across contacts with each dot showing cross-contact averaged results for an individual patient (i.e., effect size = difference between *in strength* in the interictal and *out strength* in the ictal period for each contact). Top and bottom panels show the results for the SOZ and non-SOZ contacts, respectively.

To check if this effect was also consistent at the individual contact level, we performed an additional correlation-based analysis. To that end, we evaluated the correlation between the level of interictal *in strength* and ictal *out strength* across SOZ contacts, within each individual patient (Figure 3B, top panel). Results looked like the cross-patient analysis (c.f., Figure 3A) and showed positive correlations for 9 of the connectivity measures while the rest showed negative correlations. This suggests that changes in signal characteristics and connectivity patterns from the interictal to ictal period impacts distinct connectivity measures differently. We repeated the same analysis to check the correlation between the level of interictal *out strength* and ictal *in strength* across non-SOZ contacts, which showed similar results to the SOZ contacts (Figure 3B, bottom panel). The consistent positive correlations across the nine connectivity measures support that the contacts with higher interictal *in strength* also showed a higher ictal *out strength*. However, the four measures with non-significant negative correlations, which were all frequency-domain connectivity measures, support a different transition: the contacts with higher interictal *in strength* showed a lower ictal *out strength*. This might be because of the significant changes in the frequency characteristics of signals when going from the interictal to the ictal period which continues to evolve during the seizure period (e.g., low voltage fast activity (Lagarde et al., 2019)) dominantly impacting the frequency-domain connectivity measures.

### Separation of the SOZ from the rest of the network

Having tested the “sink and source SOZ” hypotheses, we then tested to see if other node metrics would distinguish between SOZ and non-SOZ areas. These metrics were extracted from our directed connectivity measures to determine the role of each node in the network. Initially, similar to *in strength* and *out strength*, we compared SOZ and non-SOZ contacts separately for the interictal and ictal periods using four additional node metrics including *first passage time*, *clustering coefficient*, *eccentricity* and *betweenness*.

In the interictal period (Figure 4A), several node metrics discriminated SOZ from non-SOZ contacts. For instance, there was evidence (BF > 10) for higher *first passage time* in SOZ than non-SOZ for five (ANM, DI, DTF, DCOH, and PDCOH), and higher *eccentricity* for five (ANM, RECI, DI, SGC, and LMFIT) connectivity measures. *Clustering coefficient* and *betweenness centrality* showed less consistent results across connectivity measures. There was evidence (BF > 10) for higher *clustering coefficient* in SOZ than non-SOZ for three connectivity measures (ANM, DI and DCOH) but also evidence (BF > 10) for lower *clustering coefficient* in SOZ than non-SOZ for CDS. There was evidence (BF > 10) for higher *betweenness centrality* in SOZ than non-SOZ for three connectivity measures (ANM, RECI and LMFIT) but also evidence (BF > 10) for lower *betweenness centrality* in SOZ than non-SOZ for CDS. Relative consistency and non-opposing results across connectivity measures in *first passage time* suggest that SOZ areas has more complex and prolonged interactions with other areas than non-SOZ potentially due to abnormal neural activity, leading to longer *first passage times*. Higher *eccentricity* in SOZ than non-SOZ contacts suggests that the SOZ is positioned distantly from the most central nodes in the network, indicating a more peripheral position or a more complex and distributed network structure for the SOZ than the non-SOZ.

**Figure 4.**
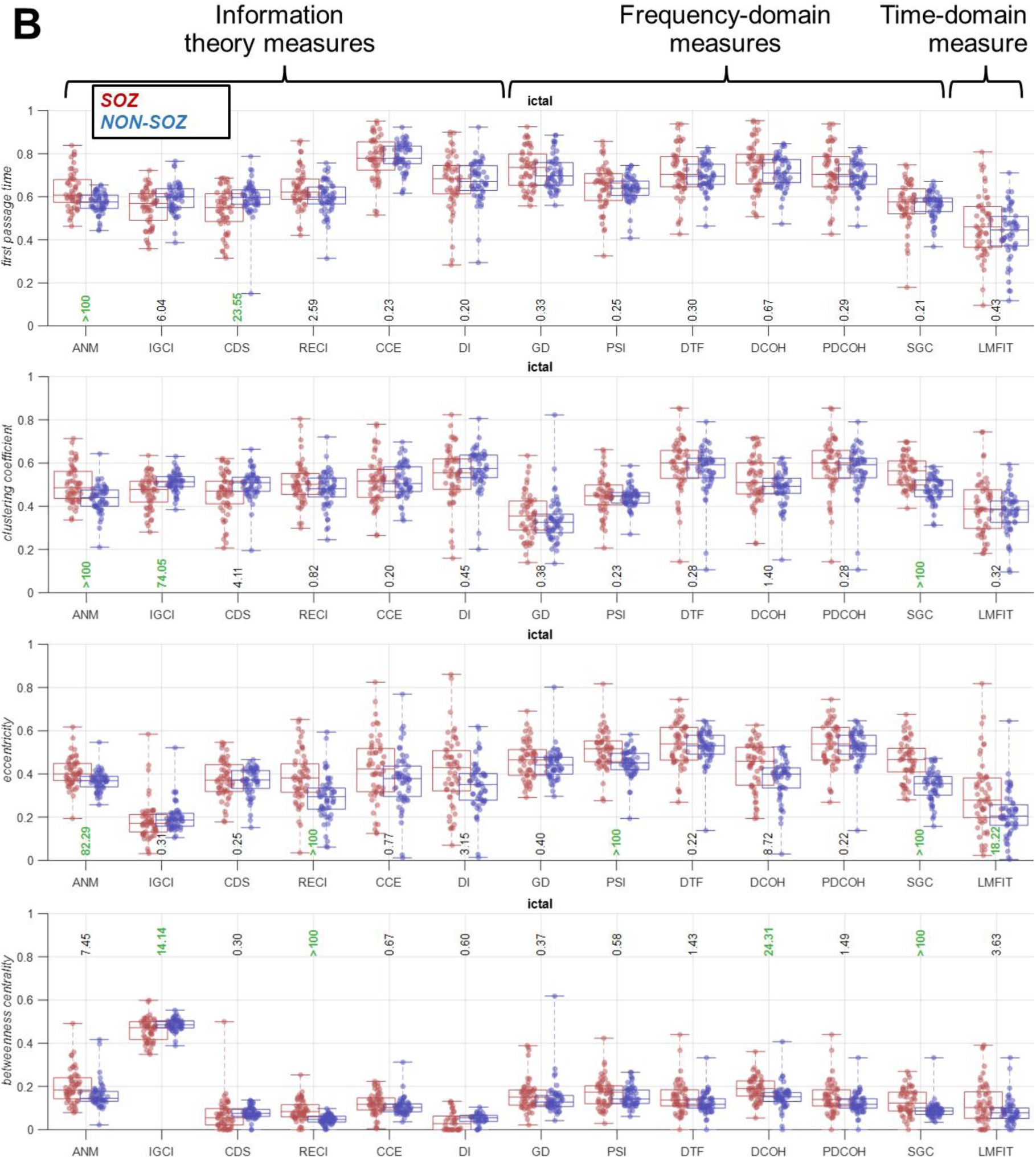
First passage time, clustering coefficient, eccentricity and betweenness centrality in the interictal (A) and the ictal (B) periods across connectivity measures. Connectivity measures are categorised into information theory, frequency-domain, and time-domain measures. Results are separated for the SOZ (red) and non-SOZ (blue) contacts with each dot showing data from one patient. Box plots show the distribution of data, its quartiles and median and whiskers indicate the maximum and minimum of the data across patients. The numbers below (above for BC) the bars indicate Bayesian evidence (BF > 10 indicated in green) for the difference between SOZ and non-SOZ data.

In the ictal period, results were less consistent across connectivity measures in terms of *first passage time* and *clustering coefficient* with two connectivity measures higher for SOZ and one higher for non-SOZ (Figure 4B). There was evidence (BF > 10) for higher *eccentricity* in SOZ than non-SOZ for five connectivity measures (ANM, RECI, PSI, SGC, and LMFIT) similar to the interictal period. There was evidence (BF > 10) for higher *betweenness centrality* in SOZ than non-SOZ for three connectivity measures (RECI, DCOH and SGC) but also evidence (BF > 10) for lower *betweenness centrality* in SOZ than non-SOZ for IGCI. These results align with the interictal results supporting that SOZ areas reside in more peripheral and less central part of the brain network reflecting their separation from the rest of the network.

### Localisation of SOZ using node network metrics

Finally, we used our set of 6 node metrics extracted from the 13 connectivity measures to see how precisely we could localise the SOZ. Localisation here refers to the discrimination of contacts within SOZ from those within non-SOZ areas. To this end, we used a decision-tree machine learning classifier to classify the SOZ and non-SOZ contacts using the 78-dimensional feature set (6 node metrics per 13 connectivity measures).

There was evidence (BF > 10) for above-chance (i.e., AUC > 0.5) localisation of the SOZ in both interictal and ictal periods at the group level (Figure 5A). At the individual level, the performance varied across patients from around chance level of 0.5 to above 0.9. These results showed that, for some patients, there was enough information in the directed connectivity measures to predict if a node was part of the SOZ or not. These results also showed that interictal activity could also provide as much localisation power as the ictal activity which is dominantly used in clinical practice.

**Figure 5.**
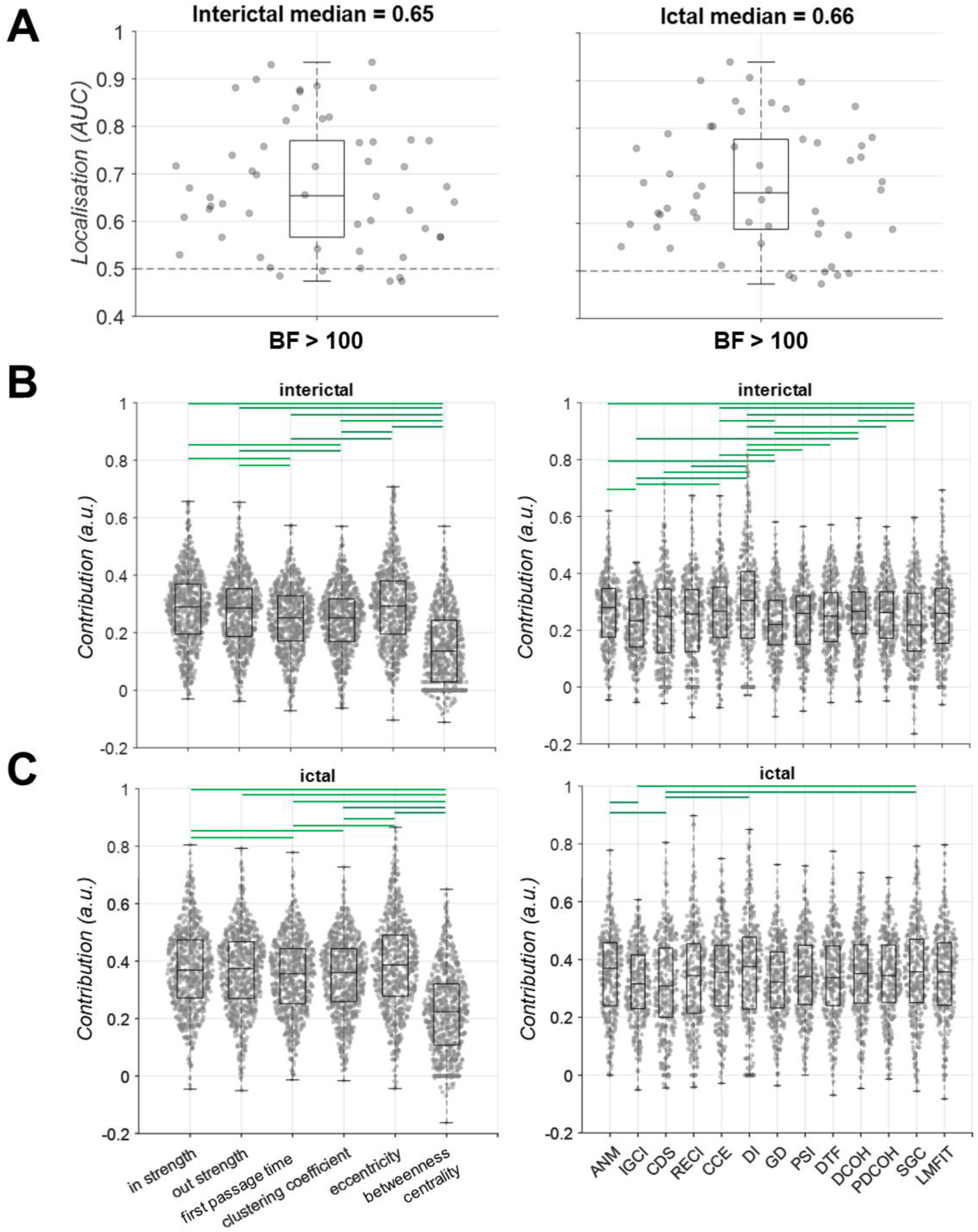
Classification of SOZ and non-SOZ contacts (localisation). **(A)** AUC of classification performance for interictal and ictal data. Box plots show the distribution of data, its quartiles and median and whiskers indicate the maximum and minimum of the data over patients. Each dot indicates the data from one patient. Numbers below the bars indicate Bayesian evidence for the difference between the true and chance performance. Horizontal dashed line refers to theoretical chance-level classification (0.5). Contribution of each connectivity measure and node metric to the classification performance shown in the interictal **(B)** and ictal **(C)** period. Contributions are calculated using random permutation in the DT classifier. Green horizontal lines indicate evidence (BF > 10) for difference between contributions.

To see if any of the patient demographic variables (i.e., surgery outcome (Engel I/Engel II-IV), region of resection (FRT/TPRMTL), pathology (lesional/non-lesional), and recording modality (SEEG/ECoG)) could explain the localisation performance, we performed a Bayes factor ANOVA t-test with these four factors as independent variables and the localisation performance (AUC) as the dependent variable. There was insufficient evidence (0.1 < BF < 10) for an effect of any of the demographic variables on the localisation performance in either interictal or ictal data (Supplementary Figure 3).

We then asked if there was systematic variation across the different recordings (i.e., time windows when the data was sampled within the interictal data and ictal/seizure data) and analysis epochs (i.e., early, mid and late). To perform this analysis, classifiers were trained and tested within each individual recording and epoch. Consistently in interictal and ictal data, we observed a higher localisation performance when localisation was done separately for each individual recording and epoch (Supplementary Figure 4) than when they were all combined (c.f., Figure 5A). Moreover, in the ictal data, we generally observed higher localisation performance in the early than mid and late epochs of data (Supplementary Figure 4B), which can be explained by a stronger separation of SOZ from non-SOZ areas at the onset of seizures. Please note that while the early epoch of data always contained the first two seconds of the ictal activity, the mid and late epochs did not match across patients as the length of seizures differed.

We then evaluated the contribution of each connectivity measure and node metric to the localisation performance. When comparing the contribution of each connectivity measure, we concatenated its node metrics and when evaluating the contribution of each node metric, we concatenated all the connectivity measures. We concatenated all patients’ data in both analyses. In the interictal period (Figure 5B), there was evidence (BF > 10) for higher contribution of *eccentricity* than *first passage time* and *clustering coefficient* and lower contribution from *betweenness centrality* than the other 5 node metrics. The median contributions of other node metrics varied in the range between the medians of *betweenness centrality* and *eccentricity*. There was evidence (BF >10) for higher contributions of DI and ANM and lower contribution of SGC than several other node metrics.

In the ictal period (Figure 5C), we observed relatively similar contributions of node metrics and connectivity measures. Specifically, node *eccentricity* and *betweenness centrality* showed the highest and lowest contributions among other node metrics. There was also evidence (BF > 10) for higher contribution of DI and ANM than other connectivity measures.

Together, these results show that *in strength* and *out strength* are informative metrics for localising the SOZ. Importantly, we observed that the node metric of *eccentricity*, which reflects how peripheral a node’s position was in the network, had even higher localisation power. We also observed that the connectivity metrics of DI and ANM provided the highest localisation power. These results were consistent across both interictal and ictal periods. We also observed that localisation was improved when done separately for each time window and was higher in early than later epochs of the ictal data. These show that, there are subtle variations in node connectivity patterns which can change the discriminability of SOZ from non-SOZ over time.

## Discussion

This work was aimed towards two goals. First, this study tested if the SOZ in people with focal epilepsy dominantly receives broadband neural activities in the interictal (resting baseline state) period (“sink SOZ” hypothesis) and dominantly transmits the activities in the ictal (seizure) period (“source SOZ” hypothesis). To that end, we utilised a data-driven approach and recruited a set of 13 directed connectivity measures along with 6 metrics of node behaviour in the network. We found that not all connectivity measures supported the above hypotheses. Nonetheless, we found evidence across several connectivity measures supporting these hypotheses. These measures showed that SOZ dominantly received neural activities in the interictal and transmitted them in the ictal period supporting the idea of seizure suppression and propagation, respectively. Second, this study evaluated the predictive power of node metrics extracted from the above-mentioned connectivity measures in localising the SOZ. To that end, we utilised the power of explainable machine learning classifiers to successfully discriminate contacts within from those outside the SOZ. This work makes several contributions to our understanding of epilepsy and how hypothesis-driven biomarkers can localise the SOZ.

Earlier studies have evaluated the directionality of signals in the interictal and ictal periods. In interictal data, some studies have suggested a leading role (higher outgoing signals) for the epileptogenic zone (EZ) (Bettus et al., 2008; Lagarde, Roehri, Lambert, Trébuchon, et al., 2018; Varotto et al., 2012) whereas others have suggested the opposite (Gunnarsdottir et al., 2022; Jiang et al., 2022; Narasimhan et al., 2020; Paulo et al., 2022; Vlachos et al., 2017). Similar discrepancy exists in studies which used ictal data, with some studies suggesting a leading role for the EZ areas (Balatskaya et al., 2020b; Courtens et al., 2016; Jung et al., 2011; Yang et al., 2018) and others providing evidence for the opposite (An et al., 2020; Janca et al., 2021; Mao et al., 2016; Nahvi et al., 2023). One important reason behind these discrepant results could be the variation in the methods used to measure directed connectivity (Doss et al., 2024; Lagarde et al., 2022; Lagarde & Bartolomei, 2024). Basically, distinct connectivity methods rely on distinct signal features to quantify connectivity. As we categorised these methods (c.f., Figure 2), some methods rely on the complexity, randomness, or the predictability of signal samples (information theory methods), whereas some rely on frequency-domain representation of signals (frequency-domain methods) and others simply rely on one-to-one mapping of time samples across areas (time-domain measures e.g., LMFIT) (Cliff et al., 2023). Even different methods within each category work differently. For example, while ANM detects causal relationship which are assumed to be additive with independent noise, IGCI can detect more complex non-additive relationships. Therefore, it is not surprising to be able to detect the directed connectivity using one method but not the other.

Building on the recent developments in neural decoding (Karimi-Rouzbahani, 2024; Karimi-Rouzbahani, Shahmohammadi, et al., 2021; Karimi-Rouzbahani & Woolgar, 2022) and connectivity analyses (Cliff et al., 2023; Karimi-Rouzbahani et al., 2022; Karimi-Rouzbahani, Ramezani, et al., 2021) and trying to avoid subjective analysis, this study adopts a data-driven and objective approach to test the direction of neural activity flow towards and away from the SOZ, which has been lacking in previous studies that tend to select a priori methods of connectivity analysis (Lagarde & Bartolomei, 2024). We showed that several connectivity measures showed higher *in strength* towards SOZ than non-SOZ areas in the interictal and higher *out strength* from SOZ than non-SOZ in the ictal period.

These results supported a switching role for the SOZ which not only supports the hypotheses of “sink SOZ” in the interictal (Gunnarsdottir et al., 2022) and “source SOZ” in the ictal period (Schindler et al., 2007), but also serves as a biomarker for localising SOZ. Interestingly, we observed a higher consistency across connectivity measures in the interictal than ictal period. This might suggest that while the inflow of activity in the interictal period might be reflected in a wider range of activity patterns as captured by a higher number of connectivity measures, the outflow of neural activity might be confined to a limited range of activity patterns (Lagarde et al., 2019). This is supported by our observation of less cross-patient generalisable epileptogenic patterns in the interictal than ictal periods (Karimi-Rouzbahani & McGonigal, 2024).

Only a few studies have evaluated directed connectivity during both interictal and ictal periods in the same patient population. For example, partial directed coherence (PDCOH) method applied to patients with type II focal cortical dysplasia showed a higher *out density* (defined as the ratio between the sum of node degrees and the total number of connections in the network) in the lesion and SOZ areas than non-SOZ areas supporting the “source SOZ” hypothesis in the ictal data, but did not find evidence to support “sink SOZ” in the interictal data (Varotto et al., 2012). On the other hand, phase transfer entropy along with node metrics applied to a sample of 43 temporal lobe epilepsy patients showed higher *out/in degree ratio* in EZ than non-EZ areas consistently through interictal and ictal data supporting “source SOZ” hypothesis in the ictal period (Wang et al., 2017). A more recent study used both interictal and ictal data and supported the “sink SOZ” hypothesis in the interictal and “source SOZ” hypothesis in the ictal data using directed transfer function (DTF) measure (Jiang et al., 2022). Finally, using a novel source-sink index obtained from both interictal and ictal activities, both the interictal sinking and ictal sourcing behaviours were observed for SOZ (Gunnarsdottir et al., 2022). In our work, DTF supported “sink SOZ” in the interictal data but showed insufficient evidence (0.1 < BF < 10) for “source SOZ” in the ictal data (c.f., Figure 2A). As these previous studies only used one (Varotto et al., 2012) or a couple (Jiang et al., 2022; Narasimhan et al., 2020; Vlachos et al., 2017) of connectivity measures, there is a possibility that they have missed some features of connectivity to comprehensively test both hypotheses. Importantly, the above-mentioned studies, which tested the directionality of connectivity did not show opposite directions to our present work (i.e., opposite directionality would mean higher *in strength* in the interictal and higher *out strength* in the ictal period for non-SOZ than SOZ). In addition to the large set of connectivity measures and node metrics evaluated in the present work, which evaluates the connectivity more exhaustively, the larger sample size used here compared to those studies allows for a more powerful evaluation.

Our results suggest that rather than being the most central/connected in the network, the SOZ seems to separate from the rest of the network. It is important to note that our measures of connectivity were extracted from temporal patterns of activity. Therefore, the separation of the SOZ from the rest of the network is more in the temporal sense than spatial and may reflect a more complex and distributed network structure for the SOZ than the non-SOZ. In other words, while spatial proximity of areas can influence the similarity of their activities, separation here means dissimilarity in activity patterns rather than spatial location. This underlines the importance of considering temporal as well spatial features when investigating epileptogenic networks (Bartolomei et al., 2017). The separation of the SOZ is consistent with previous studies which evaluated the temporal dynamics of network configurations. For example, it has been shown that, immediately after the seizure onset, the correlation in the whole-brain network drops significantly (Kerr et al., 2011; Schindler et al., 2007) possibly because of SOZ becoming functionally disconnected from other areas (Warren et al., 2010), which becomes less pronounced later in the seizure (indeed, hyper-correlation of EEG activity in the latter part of seizures has been postulated to be an emergent regulatory mechanism to promote seizure termination (Schindler et al., 2007)). This is also probably why we observed generally higher discrimination of SOZ from non-SOZ immediately after the seizure compared to later epochs (c.f., Supplementary Figure 4). It is important to note that, while our results showed evidence for neural activity dominantly flowing towards the SOZ in the interictal periods, whether this neural activity is inhibitory remains unclear. This is because connectivity methods can not determine whether the transmission is excitatory or inhibitory (Doss et al., 2024; Jiang et al., 2022; Lagarde & Bartolomei, 2024). Low voltage fast activity, the hallmark of focal seizure onset across species, has been shown to be associated with increased firing in GABA-ergic inhibitory interneurons (Gentiletti et al., 2022), triggered by accumulation of extra-cellular potassium. Studies to further evaluate links between electrophysiologic seizure evolution and associated ionic and neurotransmitter changes (e.g. using optogenetic and pharmacological approaches in animal models) have helped advance understanding of the dynamics of focal seizures (Wenzel et al., 2023), but more investigation is needed, and integrating directed connectivity methods into electrophysiologic models may be useful. Better understanding of the preictal to ictal transition may be of particular interest (Capitano et al., 2024)

Following more recent broad-band data-driven approaches (Gunnarsdottir et al., 2022), we used broad-band rather than narrow-band signals in our analyses. This aligns with studies which evaluated the connectivity over the broad-band frequency ranges and did not find any differences in directionality of signals across frequency bands (Doss et al., 2024; Jiang et al., 2022). Also, studies which suggested an effect of frequency on connectivity have reported inconsistent results. For example, while some studies have shown significantly higher *out/in degree* for EZ than non-EZ in the gamma-band activity (Wang et al., 2017) and higher outward connectivity using single-pulse electrical stimulation (Johnson et al., 2023), other studies have reported a significant decrease in outgoing connectivity from the SOZ in the gamma band frequencies during seizures (Janca et al., 2021).

Previous studies have also shown that the information in the node metrics (i.e., connectomics), extracted from directed connectivity measures, could discriminate the SOZ from non-SOZ (Sethi et al., 2016; Van Mierlo et al., 2013; Varotto et al., 2012; Vlachos et al., 2017; Wilke et al., 2011). Specifically, Wilke et al., (2011) found that the *betweenness centrality* was correlated with the location of resected cortical regions in patients with seizure-free outcomes. Van Mierlo et al. (2013) found that the electrode contacts with the highest *out degree* always lay within the resected brain regions and that the patient-specific connectivity patterns were consistent over majority of seizures. Sethi et al., (2016) analysed a network constructed from functional MRI (fMRI) data in patients with polymicrogyria and refractory epilepsy, and found that the polymicrogyric nodes showed significantly increased *clustering coefficients* and *characteristic path lengths* compared with the normal contralateral homologous cortical regions. Varotto et al., (2012) analysed the connectivity pattern in patients with type II focal cortical dysplasia and found that *out density* can discriminate SOZ from non-SOZ. Vlachos et al., (2017) evaluated effective inflow obtained from several connectivity measures including (DCOH, PDCOH and DTF) in the interictal period to show that EZ has a higher *inflow* than non-EZ. Higher *Out degree* obtained from DTF in the ictal period accurately determined the EZ nodes in (Yang et al., 2018). The present work is among the few which directly and quantitatively compared the information in several node metrics. Previously, Mao et al., 2016), who used PDC, have shown that *in degree* and *betweenness centrality* had more localisation information than *in degree* in the ictal period. In contrast, another study, which used nonlinear correlation, found more information in *out degree* than *in degree* (Courtens et al., 2016). Current study builds on these previous studies, combines a set of 6 node metrics extracted from 13 distinct connectivity measures to show how accurately they can discriminate SOZ from non-SOZ. We found evidence (BF > 10) for above-chance discrimination performance during both interictal and ictal windows, and the DI was among the most informative connectivity measures to localise the SOZ. We also found that *eccentricity* is even a more powerful biomarker for EZ localisation than *in strength* suggested in previous studies (Doss et al., 2024; Johnson et al., 2023).

It is of note that most of the previous studies have only indicated the discriminability of SOZ from non-SOZ contacts, rather than testing the generalisability of effects across new unseen contacts. Our ML-based method learns the connectivity patterns from a set of training contacts and was able to discriminate the SOZ from non-SOZ in unseen contacts. The lower performance of our node metrics, compared to our recent multi-featural localisation method on the same dataset (Karimi-Rouzbahani & McGonigal, 2024), can be explained by a variety of reasons including higher number of time windows incorporated in the analysis. It is of note that, while the results were above-chance, we did not optimise our classification/localisation pipeline, instead we focused on showing the plausibility of the method for localisation. To make the algorithm ready for real-world application, further optimisations in the pipeline can be made from the machine learning literature such as incorporating univariate signal features (Karimi-Rouzbahani & McGonigal, 2024), and data augmentation. The optimisation of the proposed pipeline is the subject of future work.

Among the 13 connectivity measures tested in this study only ANM supported both hypotheses. It may suggest that the patterns of activity and in turn connectivity significantly change from the interictal to the ictal period, which may lead to them being missed using any individual connectivity measure. More specifically, while the connectivity between brain areas might be facilitated through the modulation of signal complexities in the interictal period (as captured by CDS and DI, Figure 2A), the connectivity between brain areas might be facilitated by frequency-domain modulations in the ictal period (as captured by SGC, Figure 2B). This makes sense as ictal activities have been shown to strongly modulate the signal power in several frequency bands (Grinenko et al., 2018).

This work tested two critical hypotheses in epilepsy research and provided evidence that the SOZ seems to dominantly receive neural activities from non-SOZ potentially to be suppressed between seizures, whereas it dominantly transmits neural activities to non-SOZ during seizures. We showed that not all directed connectivity measures can detect those changes in connectivity direction from the interictal to ictal period, as probably the nature of connectivity changes with seizure onset. We also showed that, using a combination of node connectivity metrics extracted from directed connectivity measures, it is possible to localise the SOZ with above-chance performance. These results shed new light on the configuration of brain networks in epilepsy and introduces a potential method for localising the SOZ using explainable machine learning algorithms, as well as providing a rationalized set of measures for further investigation of seizure dynamics.

## Data Availability

The dataset used in this study was from previous studies and is available at https://openneuro.org/datasets/ds004100/versions/1.1.3

## Acknowledgements

We thank Mater Foundation and Mater Research Institute for supporting this study.

## Footnotes

1 In this work we define the epileptogenic zone (EZ) as the areas primarily responsible for generating seizures (Lagarde & Bartolomei, 2024; Ryvlin et al., 2024) and the seizure onset zone as where seizures start from (Bernabei et al., 2023). Nonetheless, this work analyses the directed connectivity only relative to the SOZ, but we try to use the term used in the original studies when reporting their results.

