## Supplementary material for "Directionality of neural activity in and out of the seizure onset zone in focal epilepsy"

Correspondence to: Hamid Karimi-Rouzbahani

### Supplementary material

**Supplementary table 1** Demographics of patients.

| Age | Sex | Hand | Outcome | Engel | Therapy | Implant | Resection target | Lesion status | Onset age |
|---|---|---|---|---|---|---|---|---|---|
| **40-49** | F | R | F | 3A | ABLATION | SEEG | FRONTAL | n/a | 10-19 |
| **20-29** | M | L | S | 1D | RESECTION | ECOG | FRONTAL | LESIONAL | 0-9 |
| **30-39** | M | R | S | 1B | RESECTION | ECOG | TEMPORAL | LESIONAL | 0-9 |
| **30-39** | M | L | S | 1B | RESECTION | ECOG | FRONTOPARIETAL | NON-LESIONAL | 10-19 |
| **20-29** | F | L | S | 1C | RESECTION | ECOG | TEMPORAL | LESIONAL | 0-9 |
| **50-59** | F | L | F | 4A | RESECTION | ECOG | TEMPORAL | NON-LESIONAL | 50-59 |
| **40-49** | F | L | F | 2C | RESECTION | ECOG | TEMPORAL | NON-LESIONAL | 30-39 |
| **50-59** | F | R | S | 1A | RESECTION | ECOG | TEMPORAL | LESIONAL | 30-39 |
| **20-29** | F | L | F | 2A | RESECTION | ECOG | TEMPORAL | NON-LESIONAL | 10-19 |
| **20-29** | M | L | S | 1D | RESECTION | ECOG | FRONTAL | LESIONAL | 10-19 |
| **30-39** | F | L | S | 1D | RESECTION | ECOG | TEMPORAL | LESIONAL | 0-9 |
| **20-29** | M | n/a | S | 1B | RESECTION | ECOG | TEMPORAL | LESIONAL | 20-29 |
| **40-49** | F | R | S | 1B | RESECTION | ECOG | TEMPORAL | NON-LESIONAL | 20-29 |
| **30-39** | F | n/a | S | 1D | RESECTION | ECOG | TEMPORAL | NON-LESIONAL | 30-39 |
| **30-39** | M | R | S | 1A | RESECTION | ECOG | TEMPORAL | LESIONAL | 20-29 |
| **40-49** | F | L | S | 1B | RESECTION | ECOG | TEMPORAL | NON-LESIONAL | 20-29 |
| **30-39** | M | R | S | 1A | RESECTION | ECOG | TEMPORAL | NON-LESIONAL | 0-9 |
| **40-49** | F | R | S | 1B | RESECTION | ECOG | TEMPORAL | NON-LESIONAL | 20-29 |
| **20-29** | F | R | F | 3A | ABLATION | SEEG | FRONTAL | LESIONAL | 0-9 |
| **40-49** | F | n/a | F | 3A | ABLATION | ECOG | MESIOTEMPORAL | NON-LESIONAL | 20-29 |
| **50-59** | F | R | S | 1A | ABLATION | SEEG | MESIOTEMPORAL | LESIONAL | 40-49 |
| **30-39** | M | L | S | 1A | RESECTION | SEEG | TEMPORAL | LESIONAL | 10-19 |
| **30-39** | M | n/a | S | 1A | RESECTION | ECOG | TEMPORAL | LESIONAL | 30-39 |
| **20-29** | F | n/a | S | 1A | ABLATION | ECOG | MESIOTEMPORAL | NON-LESIONAL | 22 |
| **40-49** | F | L | S | 1B | ABLATION | SEEG | MFL | NON-LESIONAL | 20-29 |
| **50-59** | F | L | F | 3A | ABLATION | SEEG | MESIOTEMPORAL | NON-LESIONAL | 40-49 |
| **30-39** | M | n/a | S | 1B | RESECTION | SEEG | FRONTAL | LESIONAL | 0-9 |
| **30-39** | M | R | F | 2A | ABLATION | SEEG | MESIOTEMPORAL | NON-LESIONAL | 30-39 |
| **30-39** | M | L | F | 4A | ABLATION | SEEG | MESIOTEMPORAL | LESIONAL | 20-29 |
| **20-29** | M | L | S | 1A | ABLATION | SEEG | PARIETAL | LESIONAL | 0-9 |
| **40-49** | F | L | S | 1B | ABLATION | SEEG | MESIOTEMPORAL | NON-LESIONAL | 20-29 |
| **30-39** | M | R | S | 1C | ABLATION | SEEG | MESIOTEMPORAL | NON-LESIONAL | 10-19 |
| **30-39** | M | n/a | S | 1D | ABLATION | SEEG | MESIOTEMPORAL | LESIONAL | 10-19 |
| **30-39** | M | n/a | S | 1D | RESECTION | SEEG | TEMPORAL | LESIONAL | 0-9 |
| **10-19** | M | n/a | S | 1A | RESECTION | SEEG | TEMPORAL | NON-LESIONAL | 0-9 |

| | | | | | | | | | |
|---|---|---|---|---|---|---|---|---|---|
| **20-29** | M | n/a | S | 1A | ABLATION | SEEG | TEMPORAL | LESIONAL | 10-19 |
| **10-19** | M | R | S | 1B | ABLATION | SEEG | INSULAR | LESIONAL | 0-9 |
| **30-39** | M | R | F | 2A | ABLATION | SEEG | MFL | NON-LESIONAL | 0-9 |
| **30-39** | M | R | F | 3A | ABLATION | SEEG | INSULAR | NON-LESIONAL | 0-9 |
| **40-49** | F | n/a | S | 1A | RESECTION | SEEG | TEMPORAL | NON-LESIONAL | 10-19 |
| **30-39** | F | n/a | F | 3A | ABLATION | SEEG | MESIOTEMPORAL | NON-LESIONAL | 10-19 |
| **40-49** | F | L | S | 1D | ABLATION | SEEG | MESIOTEMPORAL | NON-LESIONAL | 10-19 |
| **30-39** | F | L | S | 1D | ABLATION | SEEG | MESIOTEMPORAL | LESIONAL | 10-19 |
| **20-29** | M | n/a | F | 3A | RESECTION | SEEG | TEMPORAL | LESIONAL | 0-9 |
| **50-59** | M | L | F | 2A | ABLATION | SEEG | FRONTAL | NON-LESIONAL | 0-9 |
| **20-29** | F | L | F | 2A | ABLATION | SEEG | FRONTAL | NON-LESIONAL | 0-9 |
| **20-29** | F | R | S | 1A | RESECTION | SEEG | TEMPORAL | LESIONAL | 10-19 |
| **40-49** | F | R | S | 1A | RESECTION | SEEG | TEMPORAL | NON-LESIONAL | 0-9 |
| **20-29** | F | L | F | 3A | RESECTION | SEEG | FRONTAL | LESIONAL | 10-19 |
| **20-29** | F | L | S | 1A | ABLATION | SEEG | FRONTAL | LESIONAL | 0-9 |
| **30-39** | F | L | F | 3A | ABLATION | SEEG | TEMPORAL | LESIONAL | 10-19 |
| **30-39** | M | L | S | 1A | ABLATION | SEEG | MESIOTEMPORAL | LESIONAL | 0-9 |
| **20-29** | M | n/a | F | 2A | ABLATION | SEEG | MESIOTEMPORAL | NON-LESIONAL | 10-19 |
| **20-29** | F | L | F | 3A | RESECTION | SEEG | FRONTAL | LESIONAL | 0-9 |
| **20-29** | M | L | F | 3A | RESECTION | SEEG | MESIOTEMPORAL | NON-LESIONAL | 10-19 |

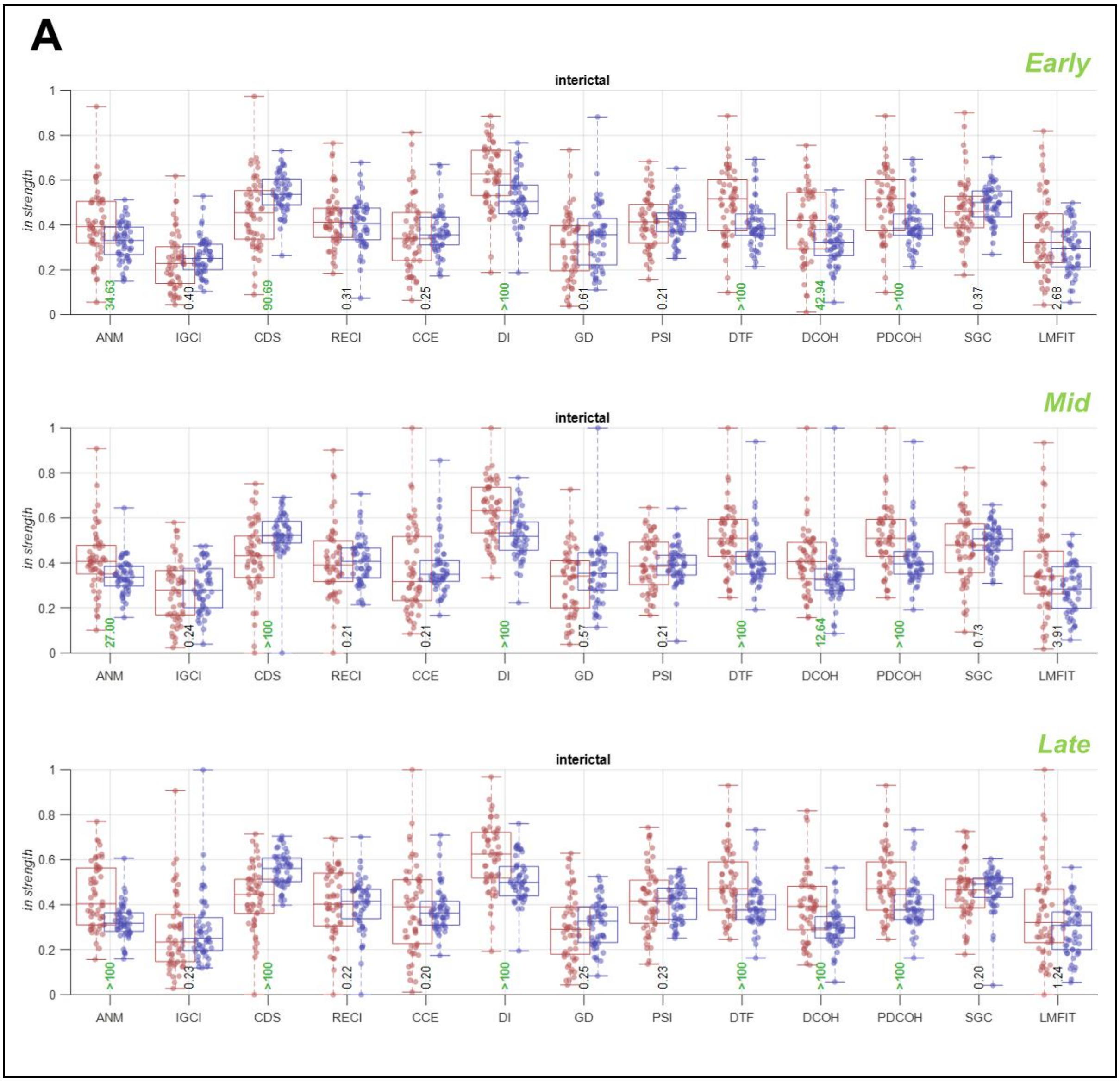

A
Early
interictal
in strength
ANM
IGCI
CDS
RECI
CCE
DI
GD
PSI
DTF
DCOH
PDCOH
SGC
LMFIT
34.63
0.40
90.69
0.31
0.25
>100
0.61
0.21
>100
42.94
>100
0.37
2.68
Mid
interictal
27.00
0.24
>100
0.21
0.21
>100
0.57
0.21
>100
12.64
>100
0.73
3.91
Late
interictal
>100
0.23
>100
0.22
0.20
>100
0.25
0.23
>100
>100
>100
0.20
1.24

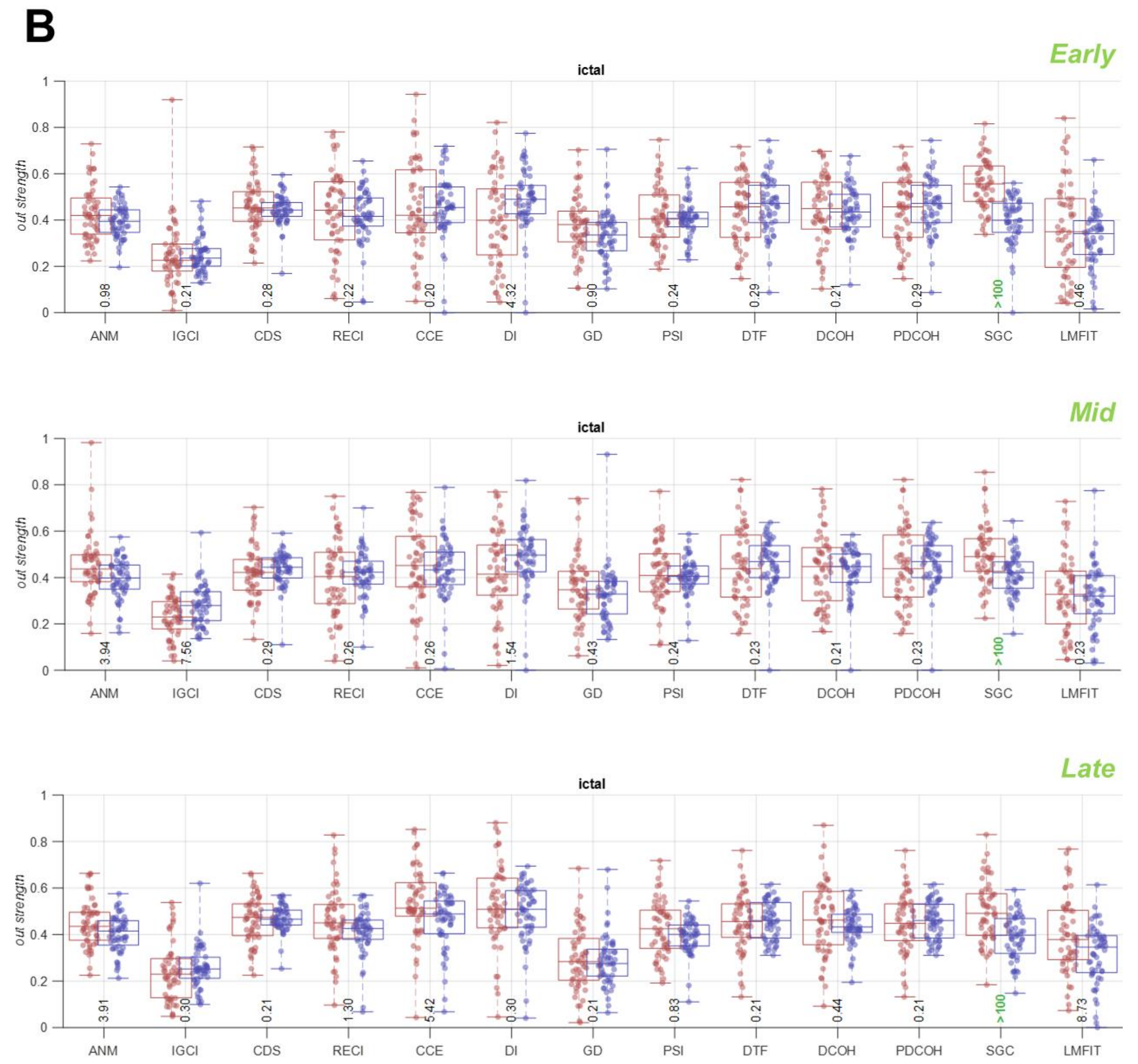


**Supplementary Figure 1 *In strength* in the interictal (A) and *out strength* in the ictal (B) period across connectivity measures using different epochs of data.** Results are separated for the SOZ (red) and non-SOZ (blue) contacts with each dot showing data from one patient. Box plots show the distribution of data, its quartiles and median and whiskers indicate the maximum and minimum of the data over patients. Numbers below the bars indicate Bayesian evidence (BF > 10 indicated in green) for the difference between SOZ and non-SOZ data.

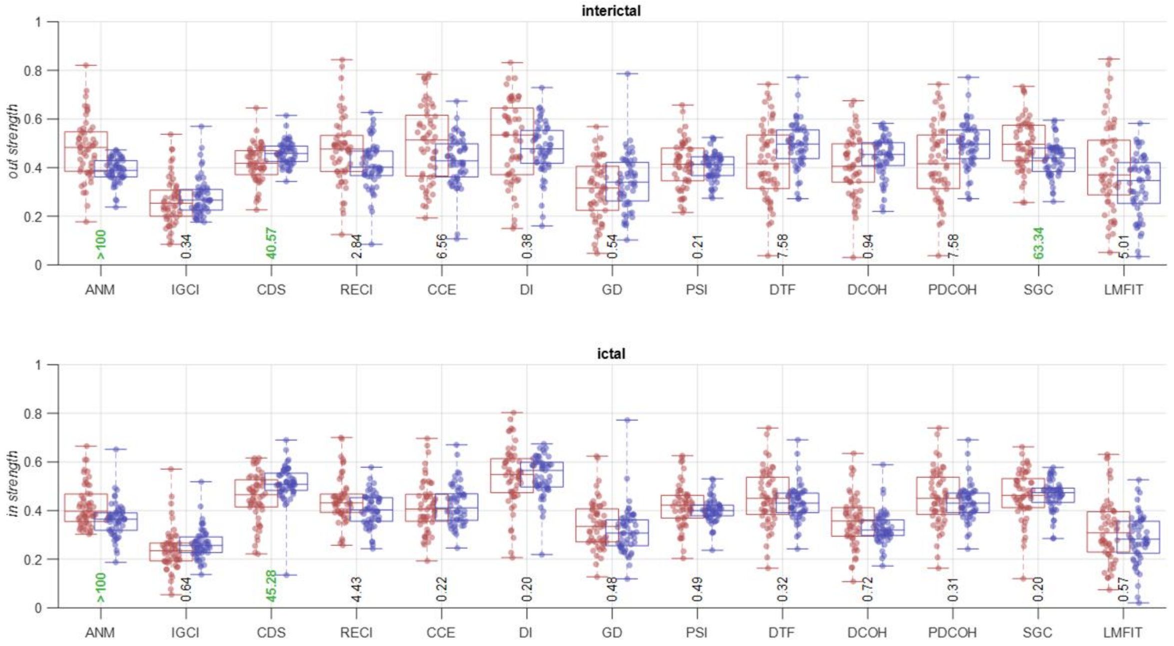


**Supplementary Figure 2** ***Out strength*** **in the interictal (top) and** ***in strength*** **in the ictal (bottom) period across connectivity measures.** Results are separated for the SOZ (red) and non-SOZ (blue) contacts with each dot showing one patient. Box plots show the distribution of data, its quartiles and median and whiskers indicate the maximum and minimum of the data over patients. Numbers below the bars indicate Bayesian evidence (BF > 10 indicated in green) for the difference between SOZ and non-SOZ data.

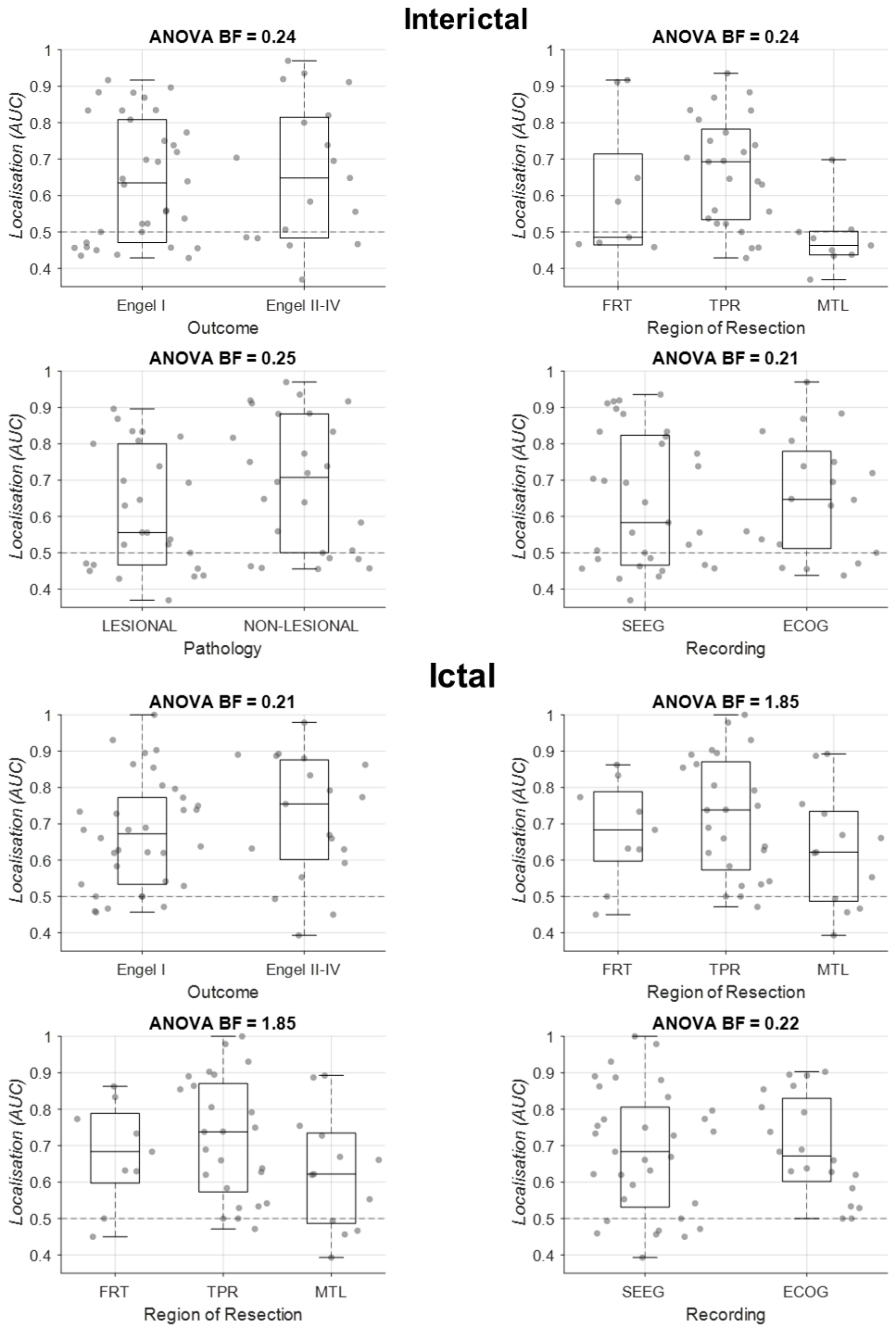


**Supplementary Figure 3** Main effects of surgery outcome (Engel I/Engel II-IV), region of resection (FRT/TPRMTL), pathology (lesional/non-lesional), and recording modality (SEEG/ECoG) on SOZ localisation. Effects were calculated using Bayes factor ANOVA. Top and bottom panels show the results for interictal and ictal classification analysis, respectively. Box plots show the distribution of data, its quartiles and median and whiskers indicate the maximum and minimum of the data across patients. Each dot indicates the data from one patient. Horizontal dashed line refers to theoretical chance-level classification (0.5). The number on the top of each panel indicates Bayesian evidence for an effect on classification performance. FRT: frontal; TPR: temporal; MTL: mesiotemporal.

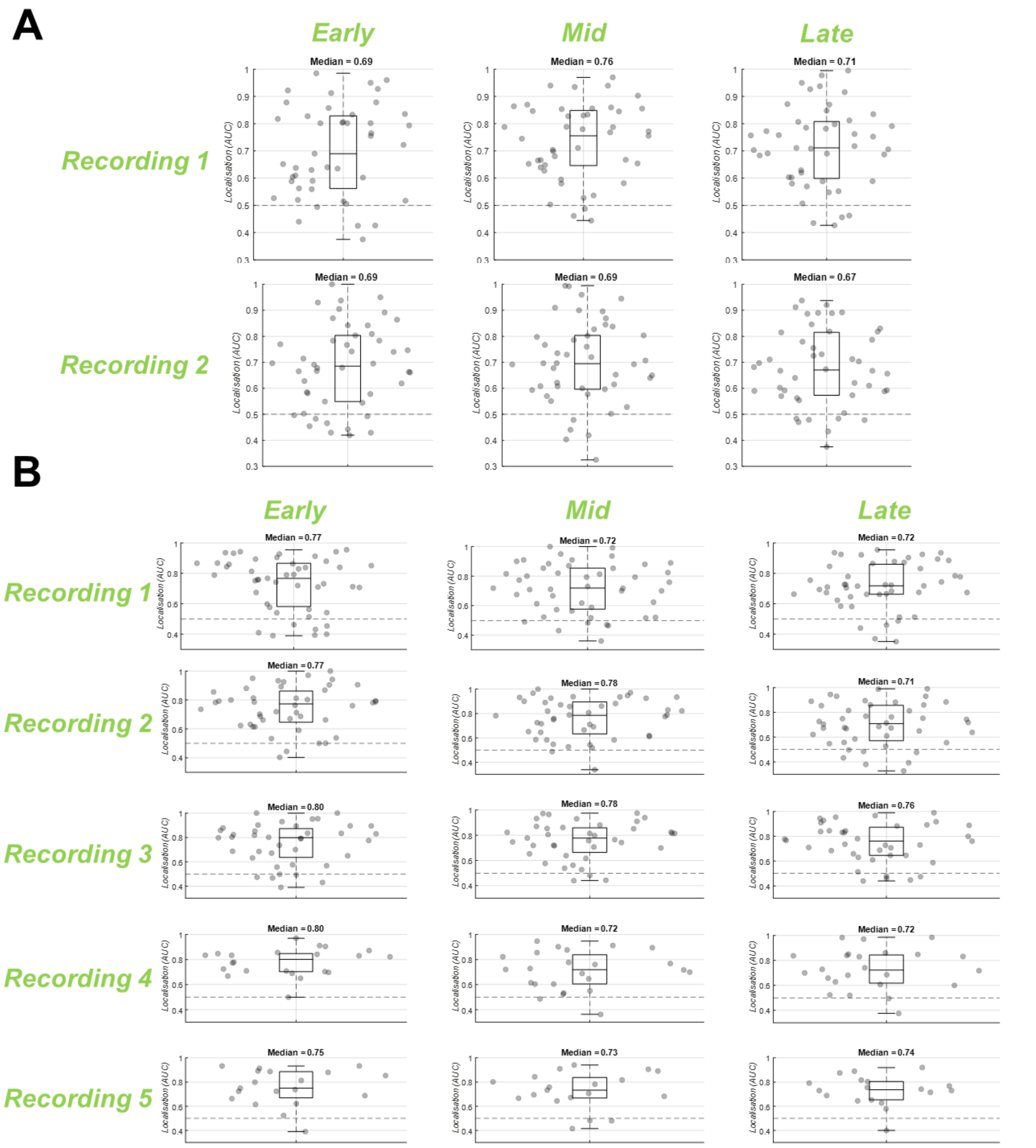


**Supplementary Figure 4 Classification of SOZ and non-SOZ contacts in different epochs of data.** AUC of classification performance for interictal **(A)** and ictal **(B)** data in different recordings and epochs. Each patient had 2 interictal recordings and 1 to 5 ictal recordings. Box plots show the distribution of data, its quartiles and median and whiskers indicate the maximum and minimum of the data over patients. Each dot indicates the data from one patient. All classification results were above chance (BF > 10; Bayesian evidence for the difference between true and null generalisation performances). Horizontal dashed line refers to theoretical chance-level classification (0.5).
